# Global Trends in Inflammatory Bowel Disease Nursing Research (2000–2024)

**DOI:** 10.1101/2025.10.12.25337837

**Authors:** Xin Zhou, Xin Zeng, Yanfei Ma, Qianmei Zhong, Qin Luo, Hong Li, Huai-li Luo

## Abstract

In this study, we examined the current landscape of global research on nursing care for inflammatory bowel disease (IBD), identified key research hotspots and emerging trends, and provided a reference for guiding future investigations. A comprehensive literature search was conducted in the Web of Science core database covering the period from January 1, 2000, to December 31, 2024. CiteSpace software was used to perform a bibliometric and visual analysis. Publication trends, country and institutional contributions, author networks, keyword co-occurrence and clustering, and citation patterns were analyzed. In total, 2,573 articles were included. The volume of publications in the field of nursing for IBD steadily increased over the past two decades. The USA produced the highest number of publications, with Brigham and Women’s Hospital—affiliated with Harvard University—leading in terms of publication output. Andrew Chan was identified as the most prolific author. *The Lancet* had the highest citation frequency. Cluster analysis revealed 15 distinct clusters, with the core research population focusing on pediatric IBD care. The cluster term “quality of life” emerged as a high-frequency keyword that remained active from 2010 to 2024 and was closely linked to chronic disease management. “Coronary heart disease” showed the strongest citation burst, highlighting a growing interest in IBD-related comorbidities. In summary, global IBD research is centered around major academic institutions such as Harvard University, with a primary focus on disease management. Emerging trends in the field include an increasing emphasis on psychosocial support, complication prevention, and comorbidities, with a progressive shift toward precision nursing.

## Introduction

With the evolution of the socioeconomic landscape and shifts in lifestyle and dietary habits, inflammatory bowel disease (IBD) has emerged as a prevalent chronic non-communicable disease worldwide. IBD encompasses a group of chronic, non-specific intestinal inflammations affecting the digestive system, with unclear etiology and pathogenesis. The two primary forms of IBD are ulcerative colitis and Crohn’s disease [1]. Common clinical manifestations include diarrhea, abdominal pain, bloody stools, mucus-containing bloody stools, and fatigue [2,3]. In recent years, the global prevalence of IBD has been increasing. The United States reports more than 1 million cases, Europe more than 2.5 million, the United Kingdom more than 500,000, and China approximately 35,000 new cases annually [4-6]. IBD is most frequently diagnosed in adults aged 18–49 years [7]. Its prolonged and recurrent course reduces the work efficiency of patients and severely affects their quality of life [8,9]. Additionally, the need for frequent long-term medical care imposes a considerable financial burden on families [10]. Research has demonstrated that effective nursing interventions can facilitate recovery and enhance the quality of life for individuals with IBD [11].

With the advancement of information technology, the nursing needs of patients with IBD have expanded. Pediatric patients with IBD have diverse nursing requirements across several domains, including physiological well-being, information support, daily life assistance, emotional and social support, and psychological well-being [12]. Therefore, it is essential to actively explore and understand the current research hotspots, trends, and shortcomings in IBD nursing to optimize care services, better address patient needs, and enhance their quality of life. Although some studies have used visualization analysis to present research hotspots and cutting-edge developments in this field, the number of documents analyzed has been relatively limited, resulting in an incomplete overview of the field [13]. CiteSpace, a widely used bibliometric visualization tool, enables the graphical representation of the literature and its interrelationships, thereby assisting researchers in identifying key topics, research frontiers, and developmental trends within a particular field [14,15]. Therefore, the present study conducted a comprehensive search of the Web of Science Core Database and applied CiteSpace software to analyze the relevant literature on IBD nursing from 2000 to 2024. This study aimed to assess global research trends in the context of IBD nursing, identify research hotspots and cutting-edge topics, and provide evidence-based insights to guide the advancement of IBD nursing practice for patients with IBD.

## Materials and Methods

### Data Sources and Retrieval Strategies

The Web of Science Core Database was selected as the data source, with the search period defined from January 1, 2000, to December 31, 2024. A combination of keyword and free-text searches was employed using the keywords “Inflammatory Bowel Disease,” “Ulcerative Colitis,” “Crohn’s Disease,” and “Nurs*.” The search was limited to documents classified as “Article” or “Review” and to those published in English, yielding a total of 2,587 records. This process is illustrated in Fig 1.

**Fig 1.**
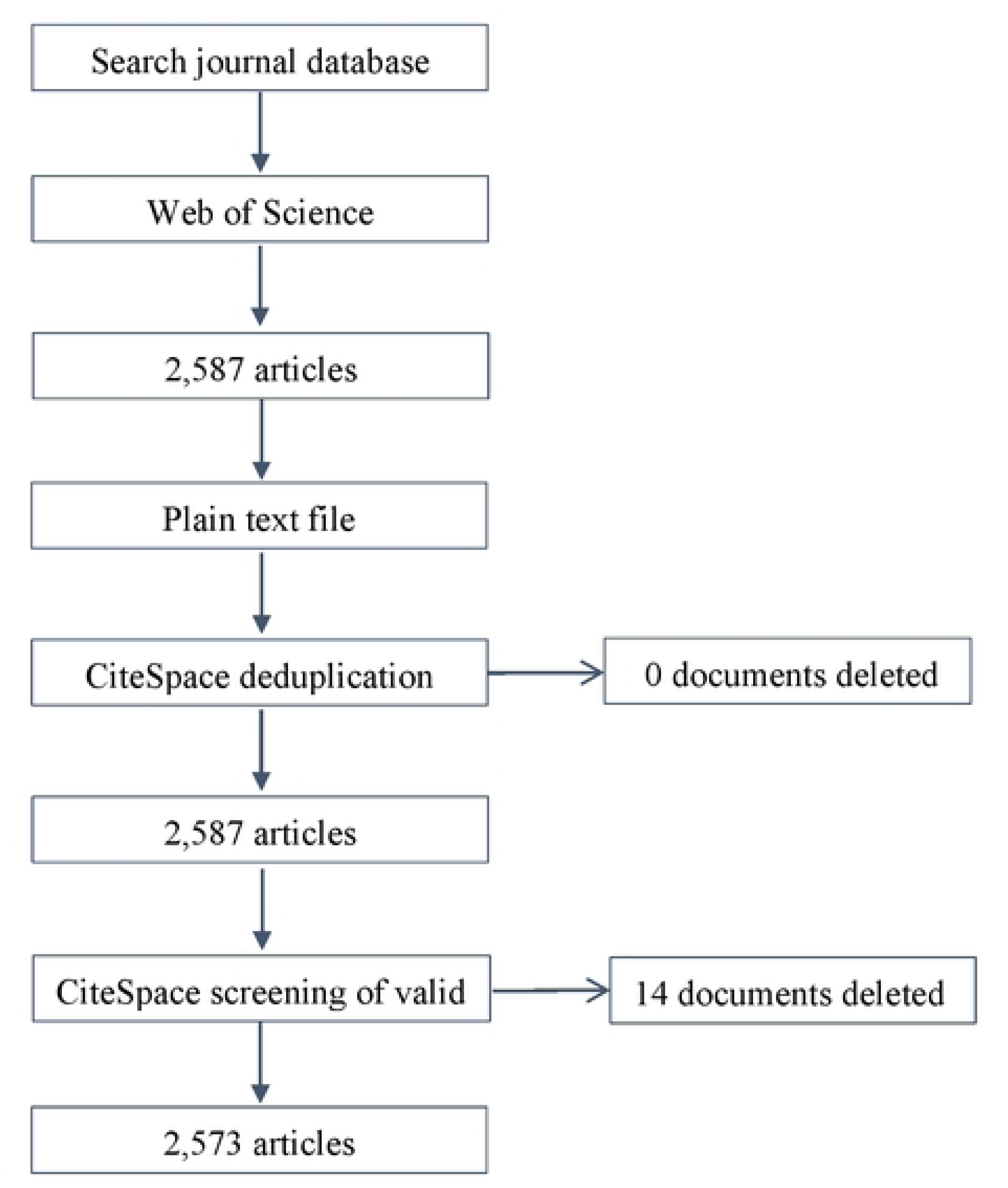
Literature search flowchart.

### Data Compilation and Analysis

Using the bibliometric visualization tool CiteSpace 6.3.R1, the search results were imported into the software in plain-text format. The time slicing was set to 1 year. The g-index was employed as the node selection criterion, with the k-value adjusted as appropriate and a standard threshold of “Top N=50.” Node types were set to country, institution, and keyword. For network pruning, “Pathfinder” and “pruning the merged network” were selected, whereas all other parameters were maintained at their default settings. WPS Office XLS was used to generate a line chart of the annual number of publications and to construct a fitted curve illustrating publication trends over time. We sorted the keywords in the published literature each year by frequency, extracted the top 10 keywords, and performed a keyword co-occurrence analysis. High-frequency keywords were clustered, and the reliability of clustering results was assessed using a modularity value (Q) >0.3 and a contour value (S) >0.5 [15]. Keyword burst detection was used to identify research hotspots within specific periods. Recent research trends were further analyzed using keyword timeline graphs. The results of keyword co-occurrence analysis, high-frequency keyword clustering, keyword salience, and keyword timeline analyses were interpreted by researchers based on their professional expertise.

## Results

### Search Results

This study retrieved a total of 2,587 articles, which were screened using the CiteSpace software, resulting in 2,573 articles being included in the final analysis.

### Current Status of IBD Nursing Research at Home and Abroad

#### Distribution of Publications

From 2000 to 2024, the global volume of research publications related to IBD nursing exhibited an upward trend. Although a slight decline was observed in 2015, the number of publications rapidly increased from 2019 to 2022, reaching a peak in 2022. Since 2020, the annual number of publications has remained consistently above 200 per year. The correlation coefficient (r) between the publication year and the publication volume was 0.928, indicating a strong positive correlation. The fitted curve equation for the annual publication volume is described by the equation y = 0.4768x² - 2.6135x + 31.511, with an R² value of 0.9467, suggesting a relatively high degree of fit. This indicates that approximately 94.67% of the variance in publication volume can be explained by the model, as illustrated in Fig 2.

**Fig 2.**
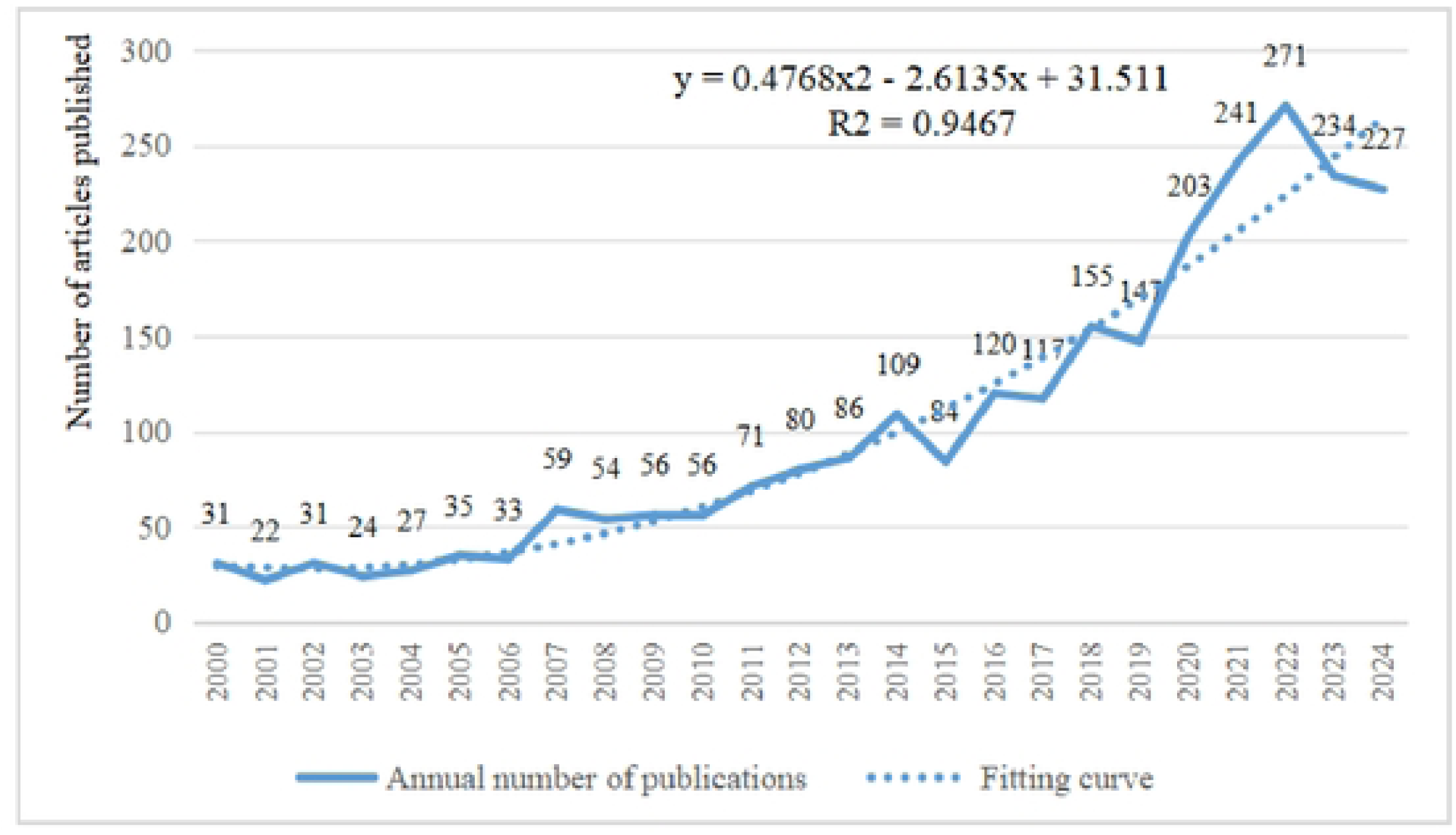
Publications in the IBD nursing literature from 2000 to 2024.

#### Distribution of Countries and Institutions Issuing Articles

Between 2000 and 2024, a total of 102 countries conducted research on IBD nursing and published articles in international journals. The United States ranked first, contributing 993 publications—accounting for 39% of the total volume—followed by China with 375 articles, representing 15%. A network visualization of country-level publication activity in the field of IBD nursing is shown in Fig 3, and the top 10 countries by publication volume are detailed in Table 1.

**Fig 3.**
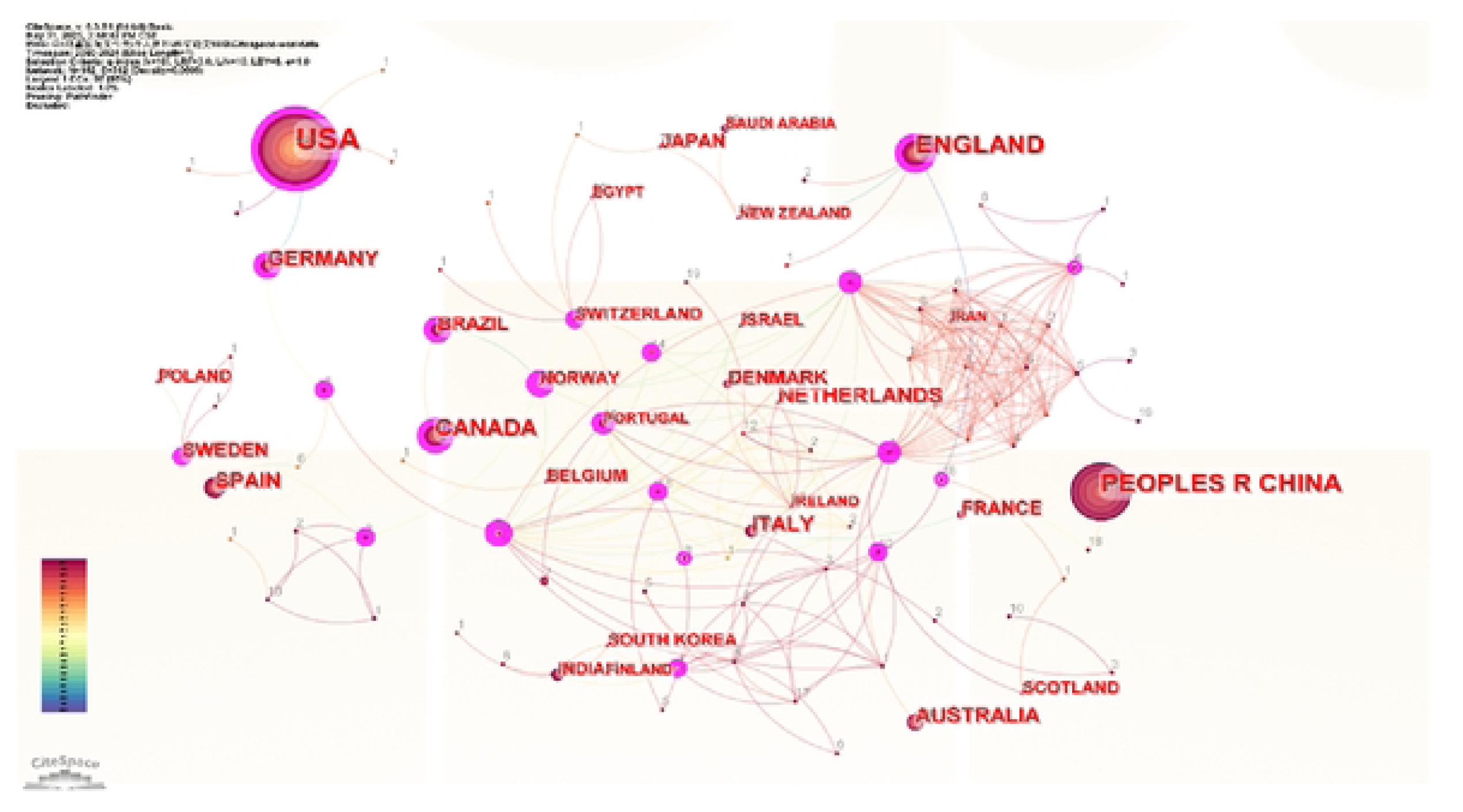
Co-occurrence map of countries with the most publications on IBD nursing from 2000 to 2024.

**Table 1.**
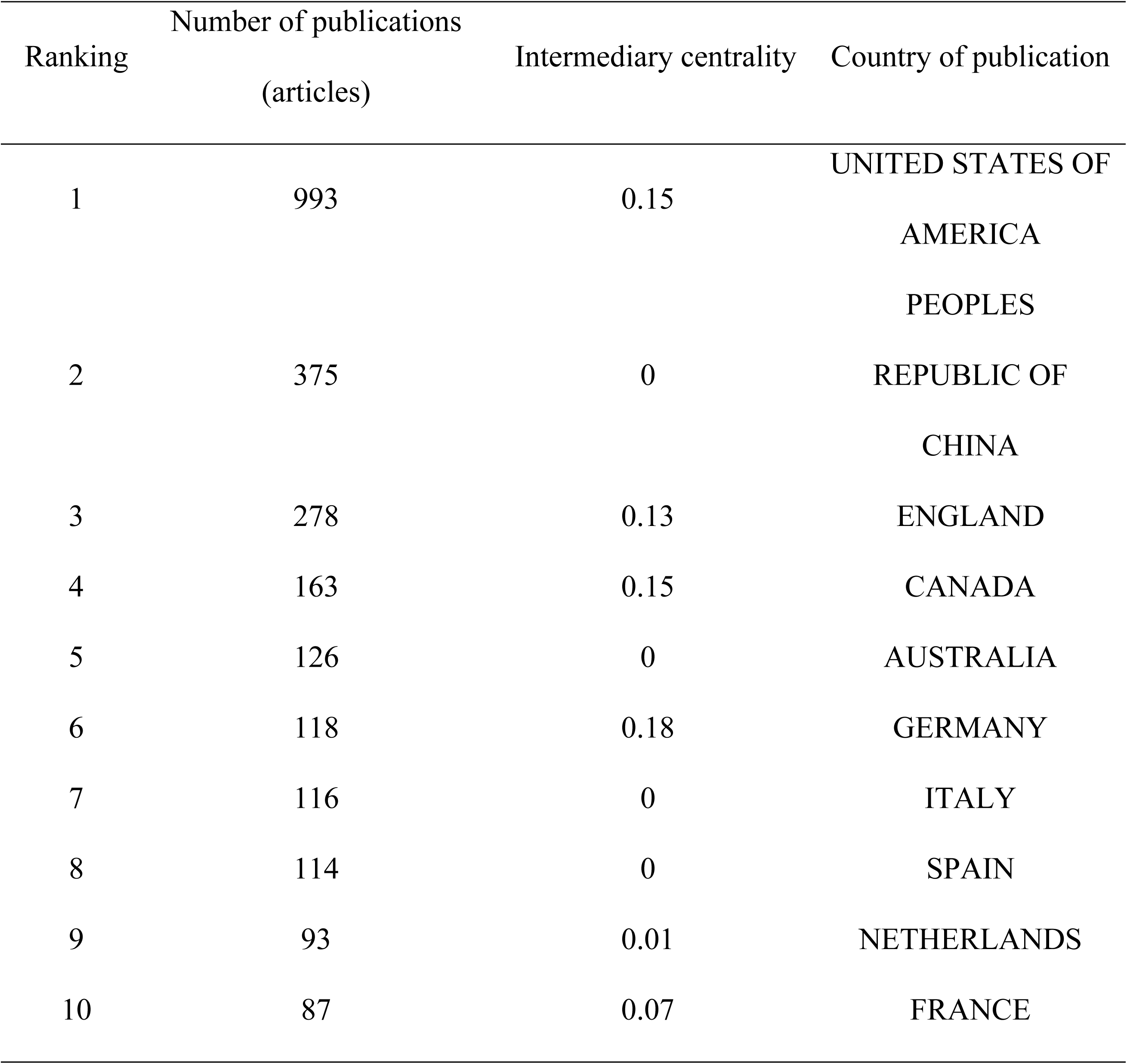
Top 10 countries with the most publications on IBD nursing from 2000 to 2024.

The top five institutions in terms of the number of publications are Brigham and Women’s Hospital (201 articles), Harvard University (142 articles), Harvard Medical School (129 articles), Massachusetts General Hospital (106 articles), and Harvard T.H. Chan School of Public Health (105 articles). The inter-institutional rankings of research institutions showed close ties and cooperation, as presented in Table 2 and shown in Fig 4.

**Fig 4.**
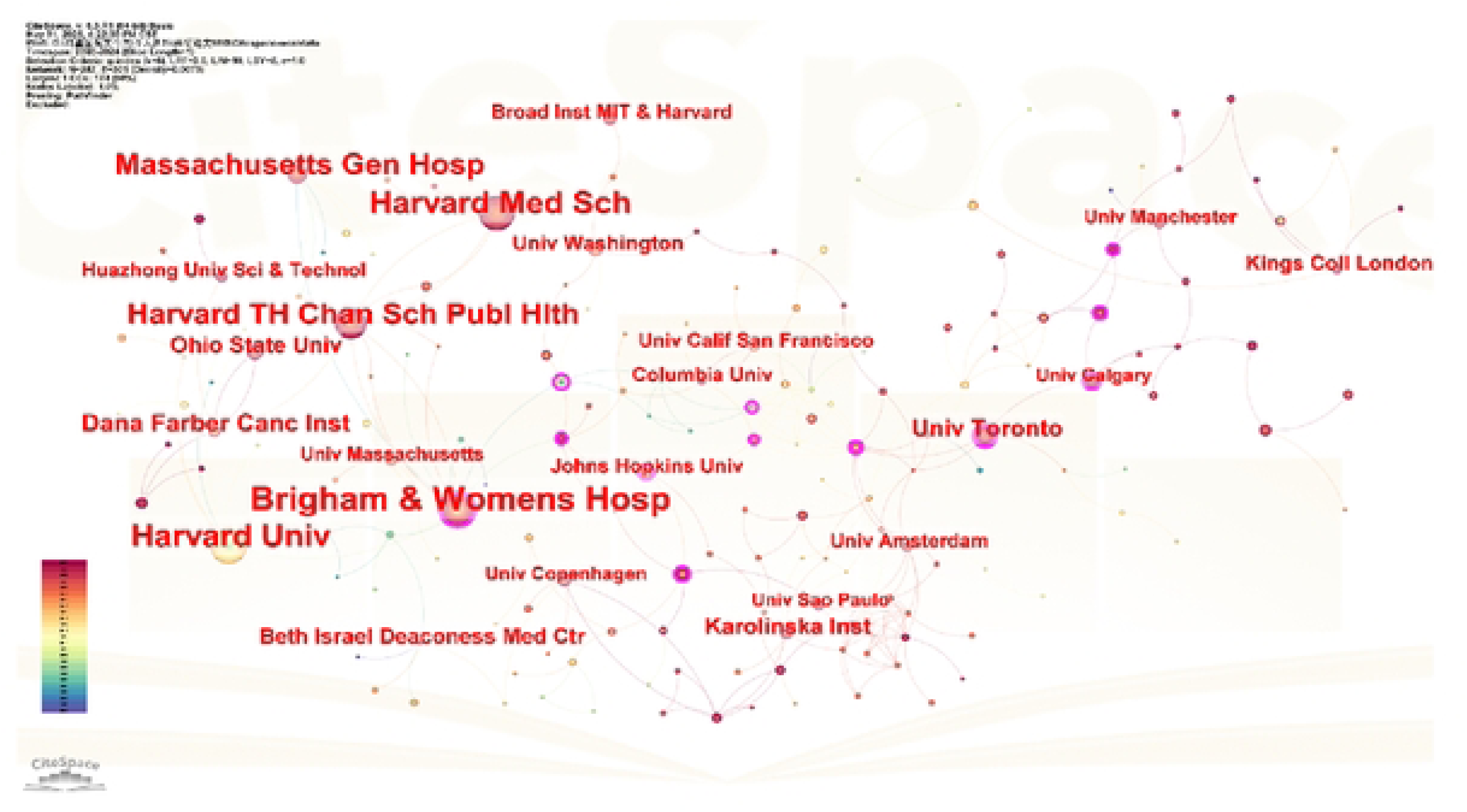
Co-occurrence map of institutions publishing in the field of IBD nursing from 2000 to 2024.

**Table 2.** Top 10 institutions publishing on IBD nursing from 2000 to 2024 ranked by centrality.

| Ranking | Number of publications (articles) | Intermediary centrality | Publishing institution |
| --- | --- | --- | --- |
| 1 | 201 | 0.32 | Brigham and Women's Hospital |
| 2 | 4 | 0.25 | Cedars Sinai Medical Center |
| 3 | 34 | 0.21 | University of Toronto |
| 4 | 15 | 0.19 | University of Calgary |
| 5 | 20 | 0.17 | Johns Hopkins University |
| 6 | 8 | 0.17 | Case Western Reserve University |
| 7 | 13 | 0.12 | Children's Hospital |
| 8 | 7 | 0.12 | University of Sydney |
| 9 | 4 | 0.11 | University of Chicago |
| 10 | 4 | 0.11 | Leeds Teaching Hospitals NHS Trust |

The top five authors in terms of the number of publications are Andrew T. Chan (70 articles), Charles S. Fuchs (37 articles), Edward L. Giovannucci (34 articles), Frank B. Hu (34 articles), and Ashwin N. Ananthakrishnan (34 articles), as shown in Fig 5.

**Fig 5.**
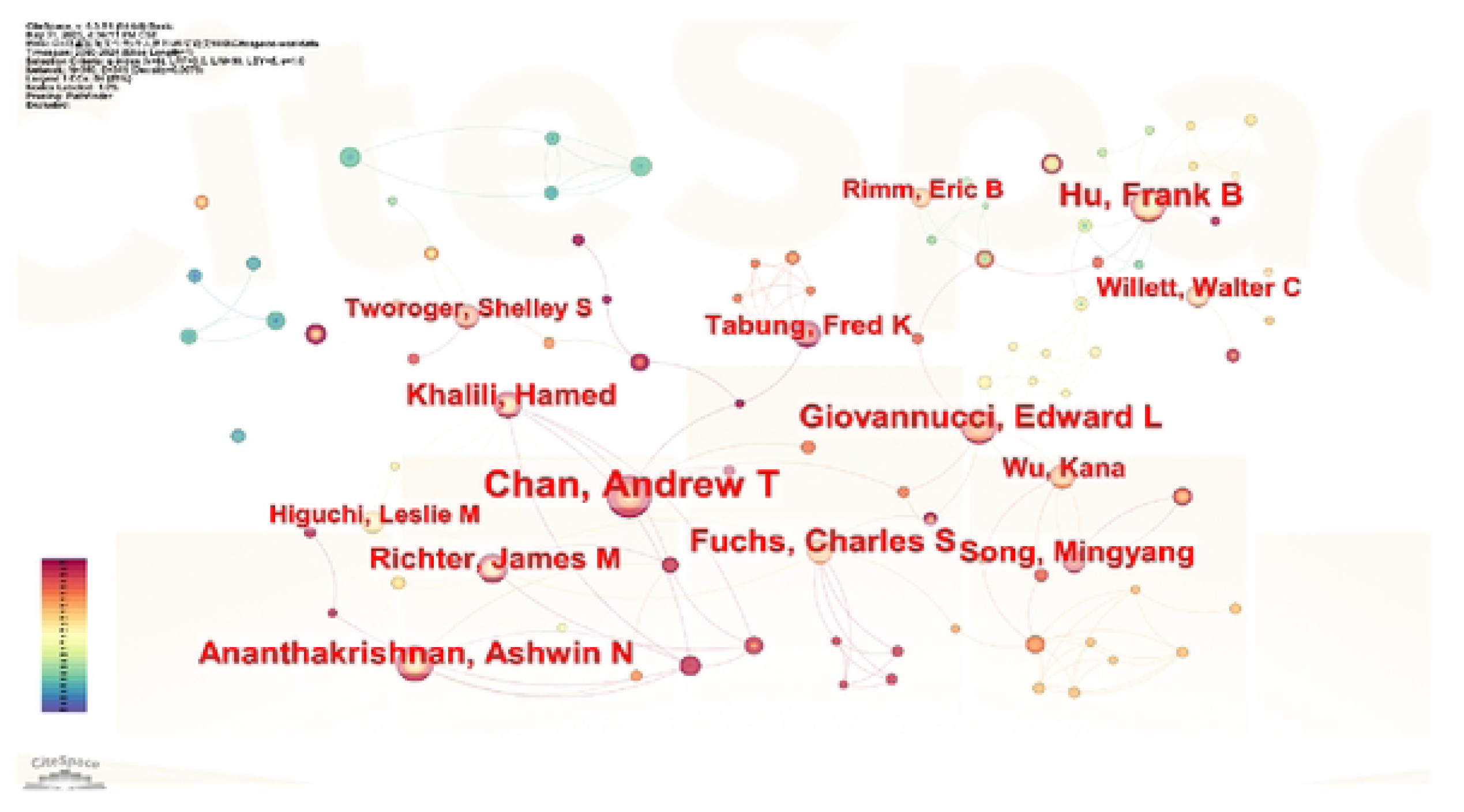
Co-occurrence map of authors in the field of IBD nursing published between 2000 and 2024.

The top five journals in terms of citations are *Lancet* (819 times), *New England Journal of Medicine* (766 times), *Journal of the American Medical Association* (632 times), PLOS ONE (583 times), and Gastroenterology (479 times), as shown in Fig 6.

**Fig 6.**
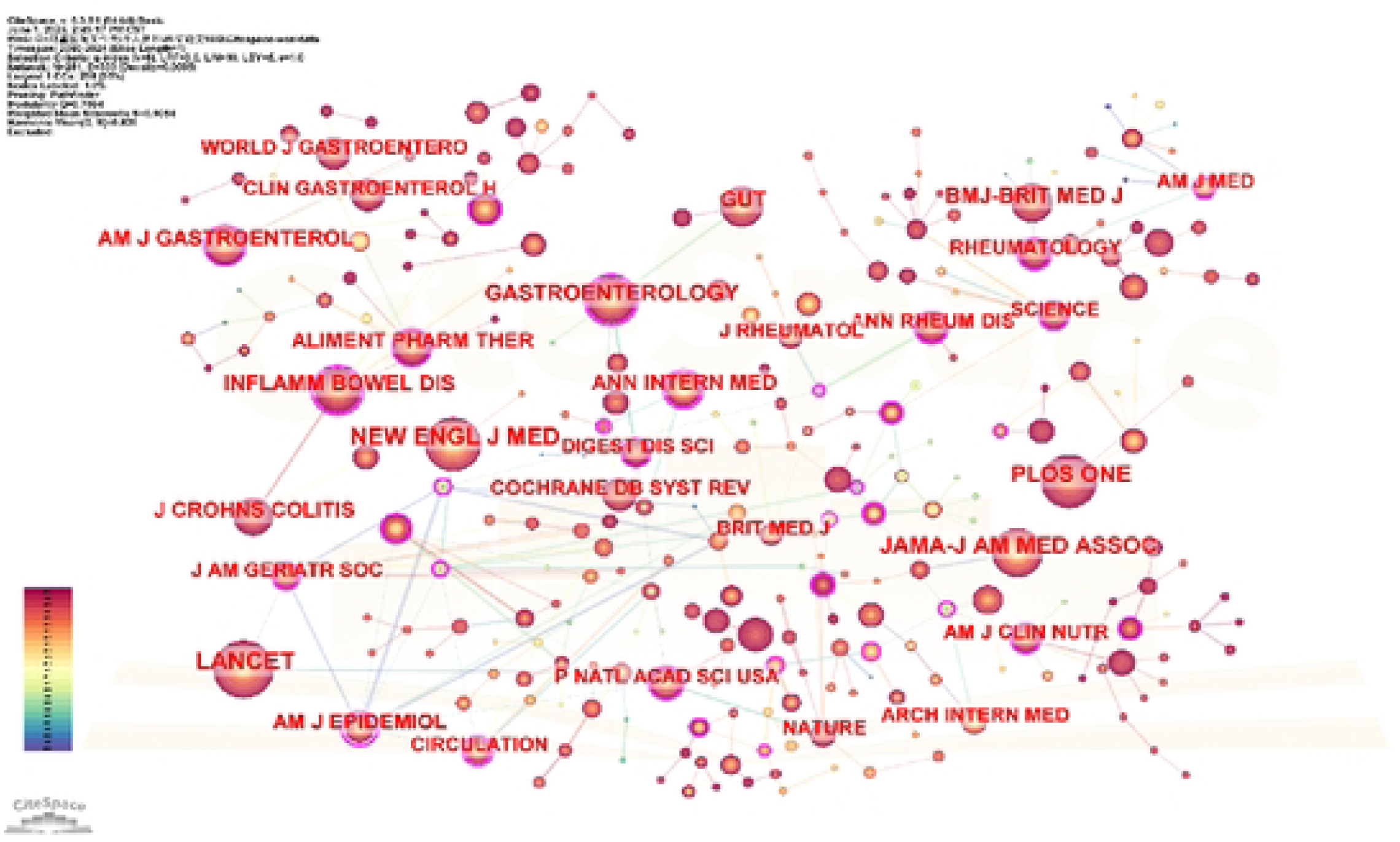
Co-occurrence map of journals cited in the field of IBD nursing from 2000 to 2024.

### Research Hotspots in IBD Nursing

#### Keyword Co-occurrence Analysis

To identify the research hotspots in IBD nursing, keywords from the published literature were ranked annually by frequency, and a keyword co-occurrence analysis was performed. The resulting keyword co-occurrence map is presented in Fig 7, and the top 10 keywords by co-occurrence frequency are listed in Table 3. Among these, the most important keywords were “management,” “quality of life,” and “risk.” Management primarily includes symptom management, nursing management, and clinical management guidelines, whereas risk includes the risk of disease onset and the development of complications.

**Fig 7.**
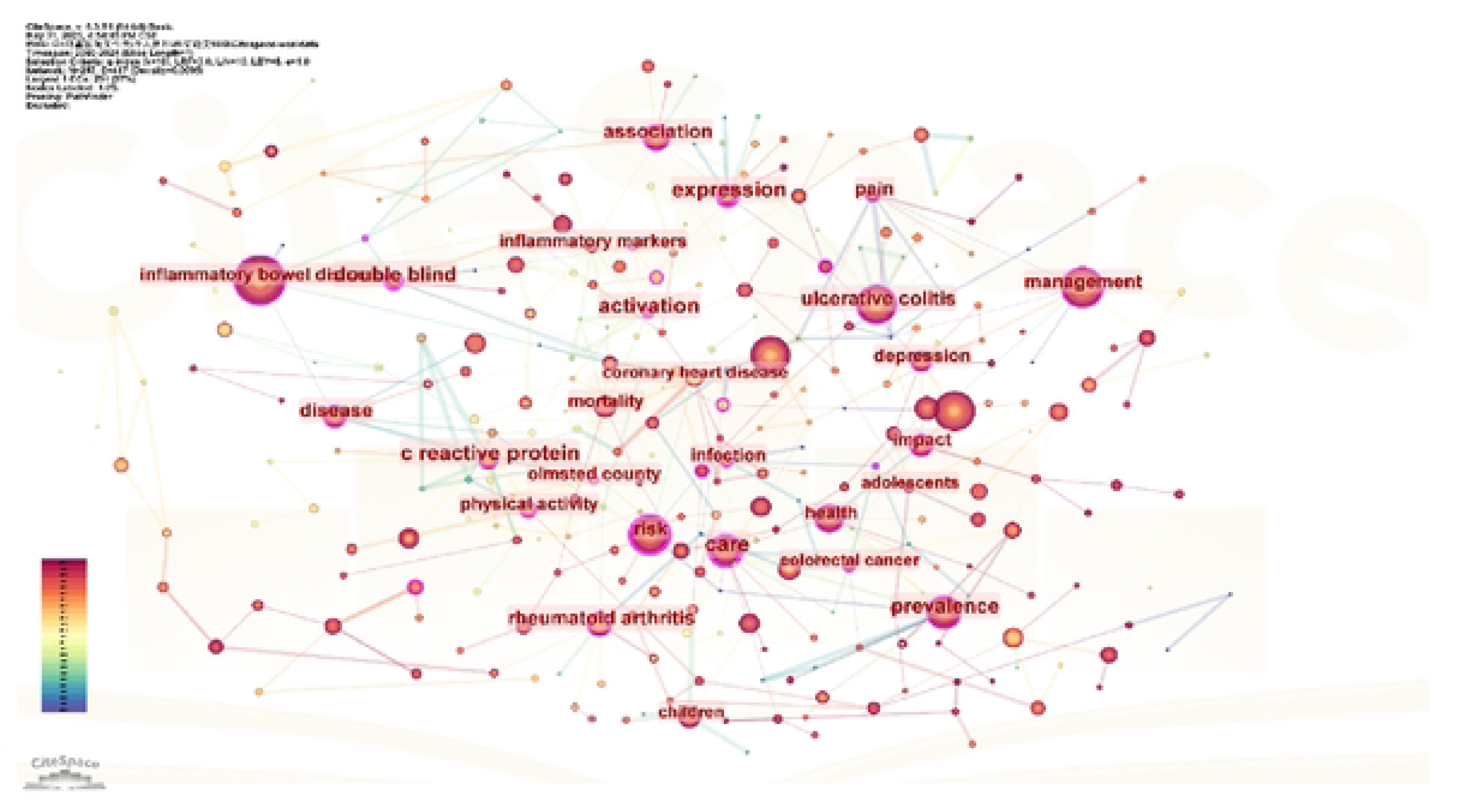
Co-occurrence map of keywords in the field of IBD nursing from 2000 to 2024.

**Table 3.**
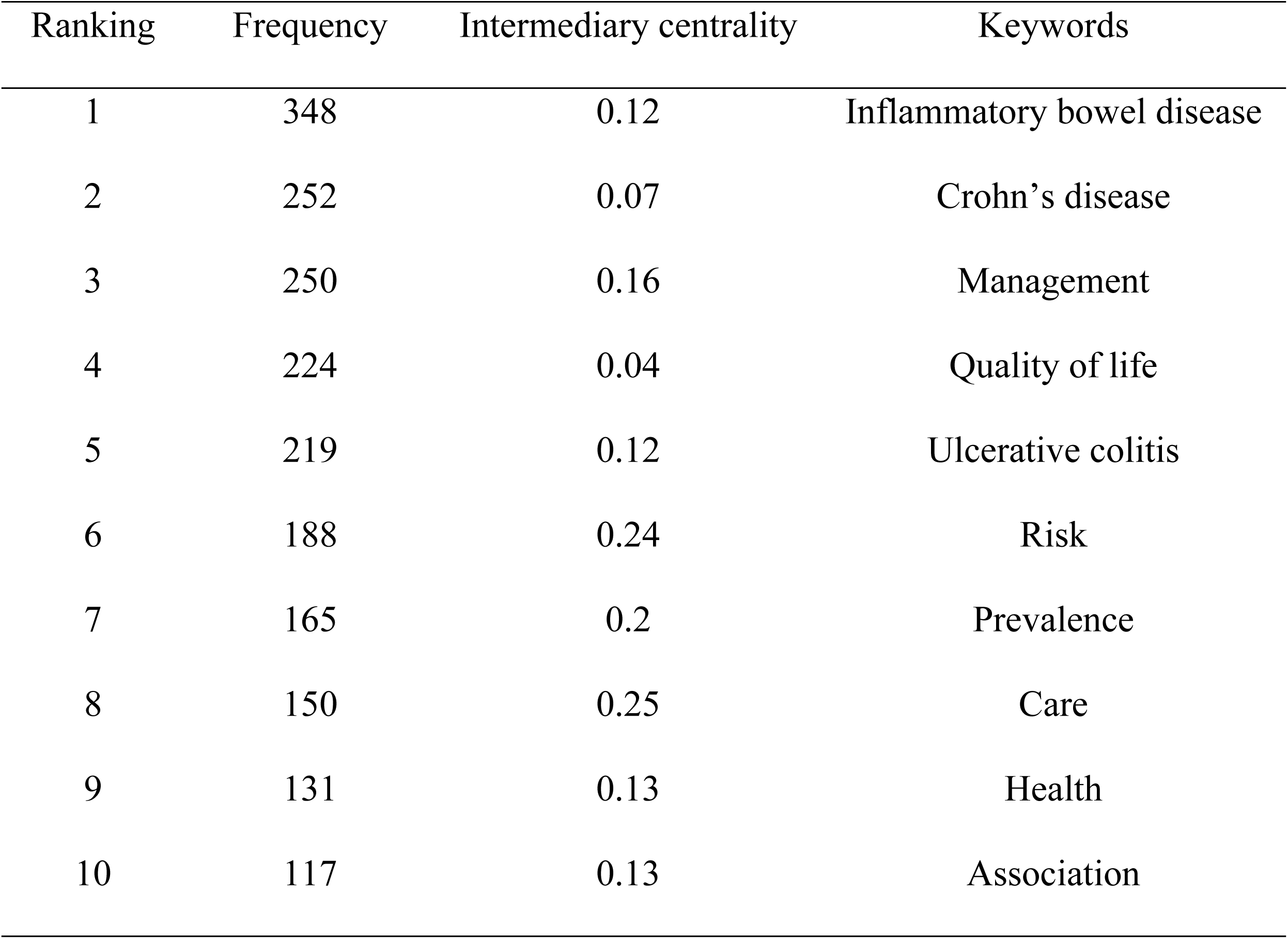
Top 10 co-occurrence frequencies of keywords in IBD nursing from 2000 to 2024.

| Ranking | Frequency | Intermediary centrality | Keywords |
| --- | --- | --- | --- |
| 1 | 348 | 0.12 | Inflammatory bowel disease |
| 2 | 252 | 0.07 | Crohn's disease |
| 3 | 250 | 0.16 | Management |
| 4 | 224 | 0.04 | Quality of life |
| 5 | 219 | 0.12 | Ulcerative colitis |
| 6 | 188 | 0.24 | Risk |
| 7 | 165 | 0.2 | Prevalence |
| 8 | 150 | 0.25 | Care |
| 9 | 131 | 0.13 | Health |
| 10 | 117 | 0.13 | Association |

#### Keyword Cluster Analysis

Keyword cluster analysis produced 15 distinct clusters, as shown in Fig 8. Clusters were ranked by the number of keywords they contained, with labels generated using the log-likelihood ratio algorithm. The cluster number is inversely proportional to the cluster size; that is, a smaller cluster number corresponds to a larger cluster size and a greater number of keywords (size value) [15]. In this study, the results of the keyword clustering analysis yielded a Q value of 0.7594 and an S value of 0.9054, indicating that the clustering results are highly reliable.

**Fig 8.**
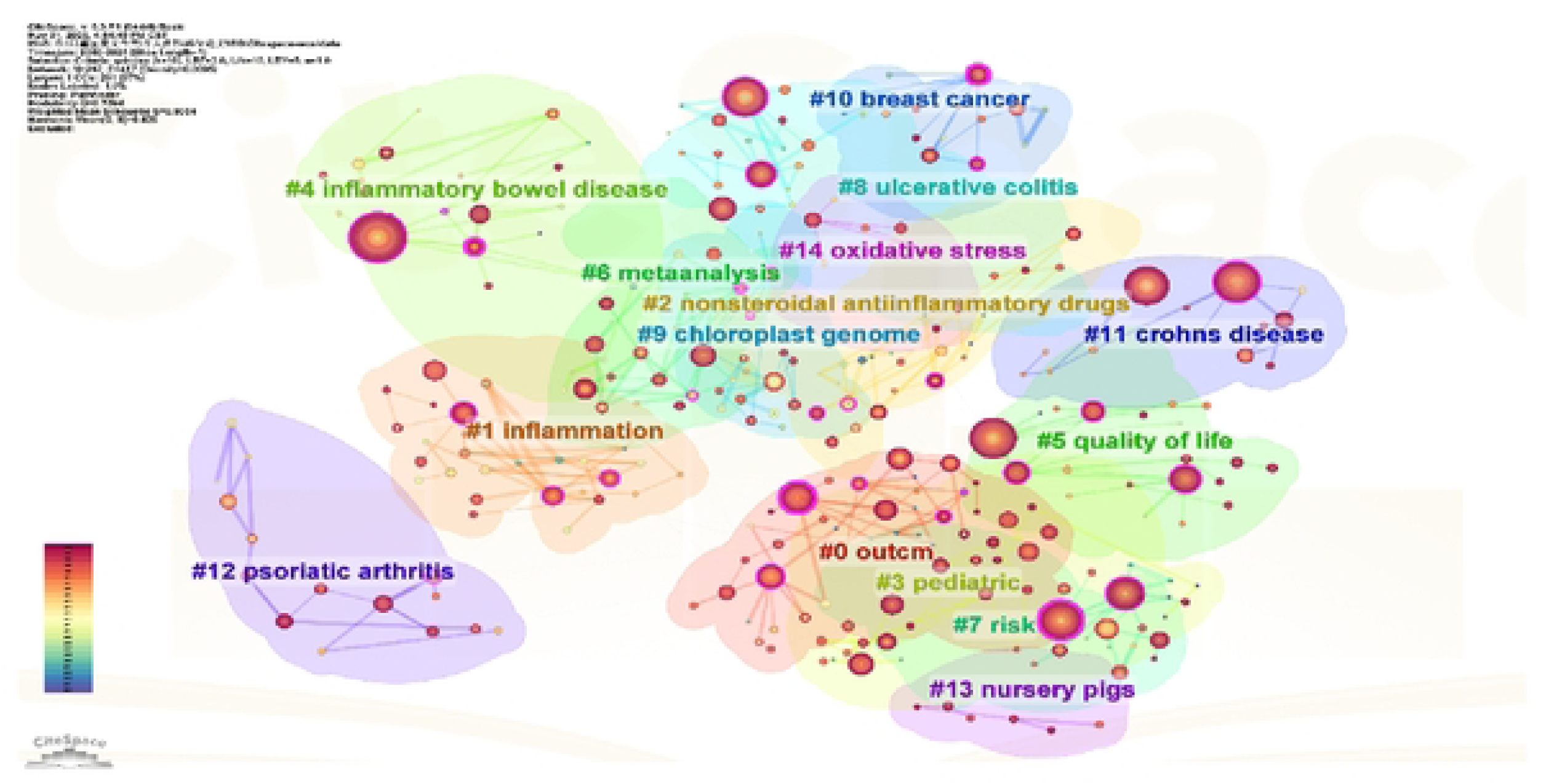
Clustering diagram of keywords in the field of IBD nursing from 2000 to 2024.

#### Keyword Salience Analysis

Using the software’s burst detection function, keyword burst analysis can predict research hotspots in a particular field during specific periods. “Begin” indicates when a keyword begins to burst, “End” represents when it stops, and “Strength” reflects the intensity of its use during that period [16]. This study showed that the highest strength value was for coronary heart disease, at 11.43. Between 2000 and 2010, the research primarily focused on drug therapy, biological mechanisms, and cardiovascular comorbidities. Research methodologies gained prominence from 2010 to 2020, with primary emphasis on randomized controlled trials and meta-analyses. Since 2020, research has shifted toward precision medicine and specialized nursing care, focusing on topics such as for COVID-19, quality of life, gut microbiota, and efficacy, as shown in Fig 9.

**Fig 9.**
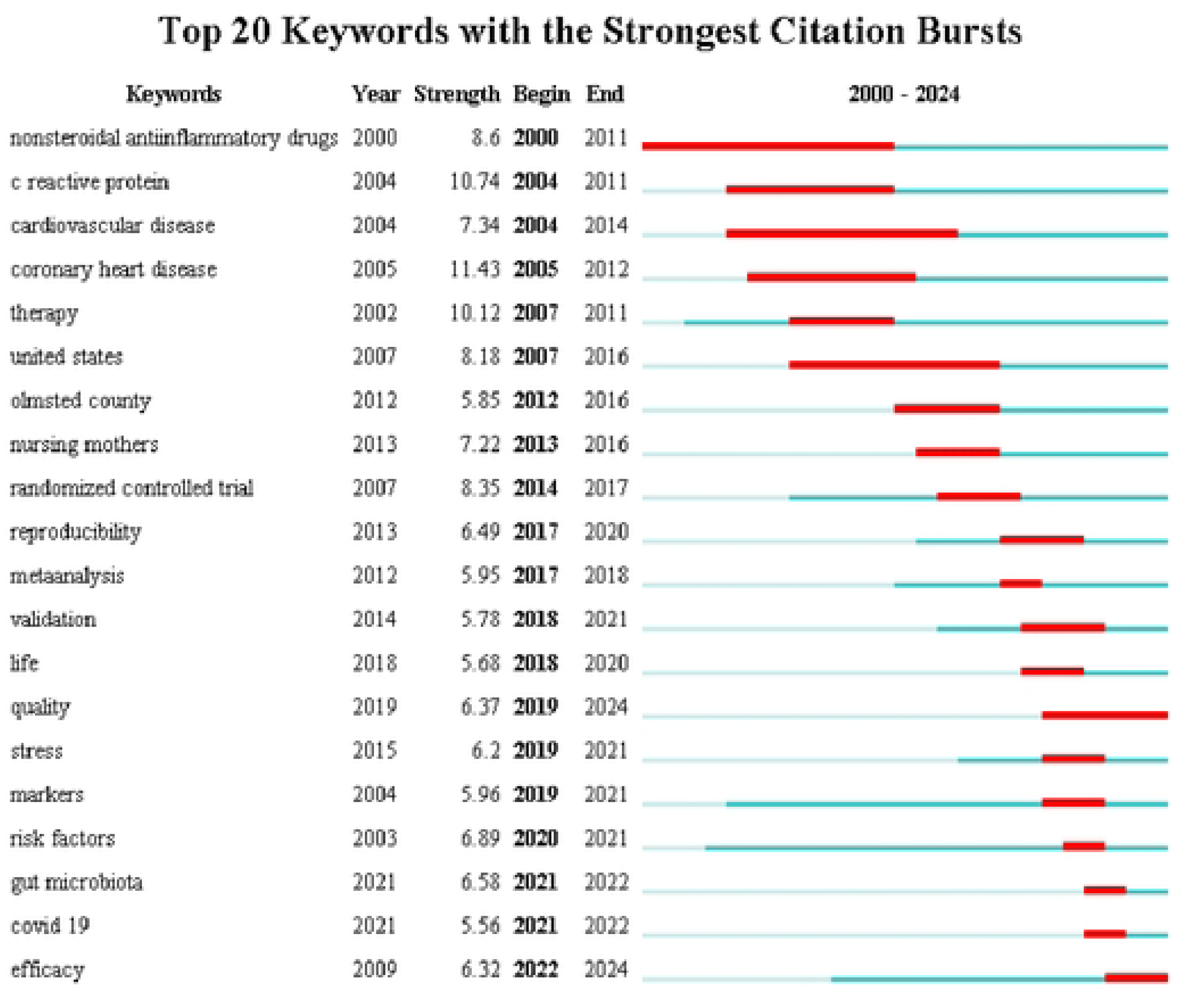
Keyword emergence map of IBD nursing literature from 2000 to 2024.

#### Keyword Timeline Analysis

In recent years, global research on IBD has primarily concentrated on clinical outcomes, non-steroidal anti-inflammatory drugs, quality of life, and disease risk prediction. Core research efforts have largely focused on the care of pediatric IBD patients. The keyword “quality of life” remained consistently active from 2010 to 2024 and was closely associated with the chronic management of IBD, as shown in Fig 10.

**Fig 10.**
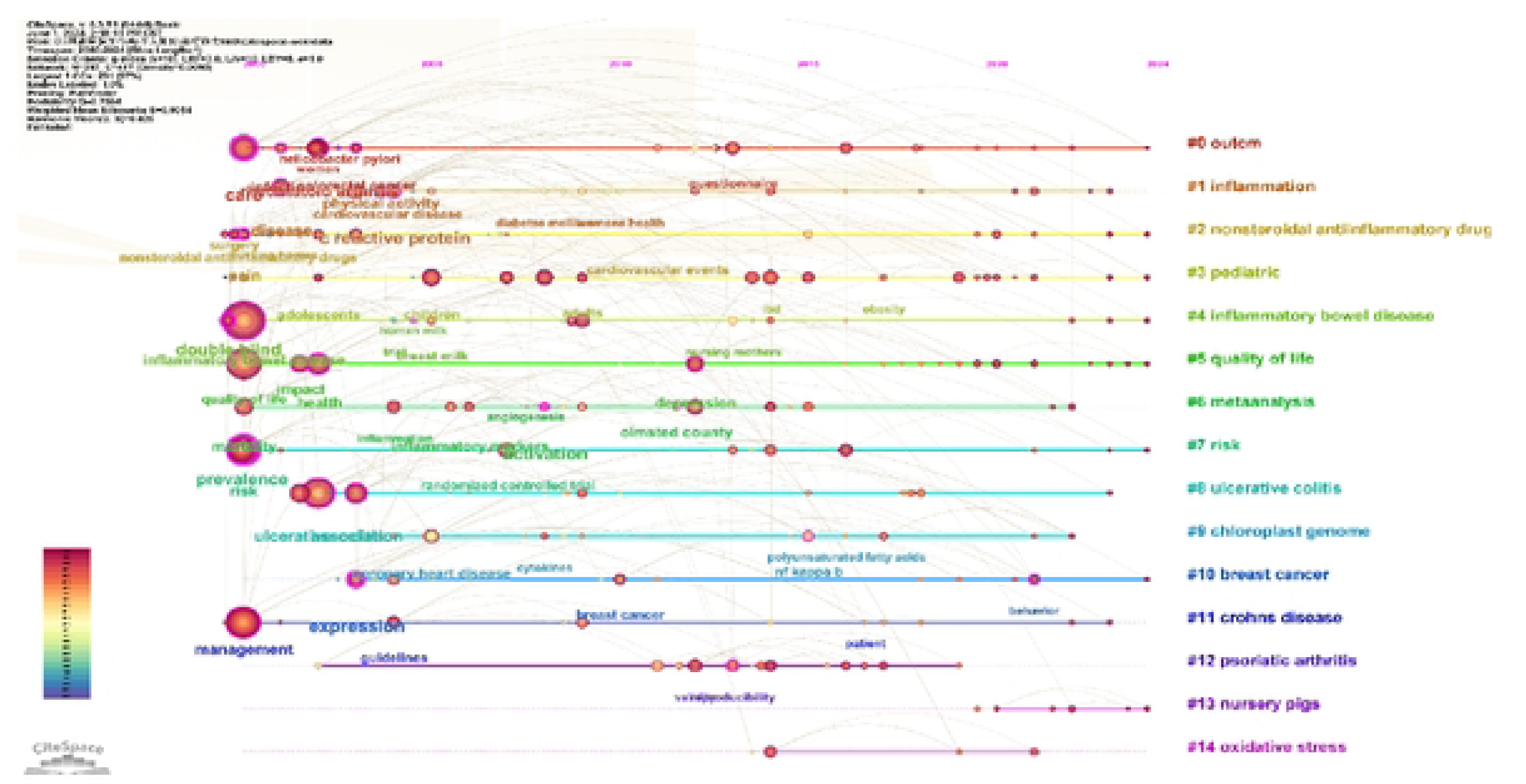
Timeline map of keywords related to IBD nursing literature from 2000 to 2024.

## Discussion

### Analysis of the Current Status of Global IBD Nursing Research

#### Publication Volume Analysis

Research indicates that IBD care studies have steadily increased over time, reflecting growing interest and maturation of the field. The R^2^ value of the fitted curve was 0.9467, further supporting the projection that IBD care will continue to be a significant focus of future research. With the advancement of information technology and the implementation of supportive national policies [17], there is a growing demand for intelligent self-management among patients with IBD [18], along with the implementation of continuous care for patients with IBD based on information technology [19]. As a result, this field is expected to continue to garner substantial scholarly attention.

#### Analysis of Issuing Countries and Institutions

The results indicated that the United States produced the highest number of publications in the field of IBD nursing, followed by China. Among the contributing institutions, Brigham and Women’s Hospital had the largest number of publications, forming a research cluster centered around the Harvard University system in the Boston area of the United States. Notably, the top five institutions by publication volume were found to be located within this region. Brigham and Women’s Hospital ranked first in publication volume and also had the highest intermediary center score (0.32), indicating that it has conducted in-depth research in IBD care and serves as the most critical hub within the research collaboration network. It maintains numerous cooperative relationships with other research institutions. Overall, the institutions involved in IBD nursing research have established a wide-reaching global collaboration network, prominently represented by North American institutions, with additional significant contributions from Europe, Australia, and other regions. Within this network, teaching hospitals and medical centers—such as Brigham and Women’s Hospital and Massachusetts General Hospital—occupied central positions, indicating that IBD care is primarily rooted in clinical practice. Most leading research institutions in this field are healthcare facilities that provide direct patient care.

#### Author Analysis

The results indicated that research in the field of IBD care is primarily centered around scholars affiliated with Harvard University in the United States (especially Andrew T. Chan and Edward L. Giovannucci), maintaining a close collaborative relationship. Edward L. Giovannucci functions as a key hub within the collaborative network, whereas Andrew T. Chan is the most prolific author, exerting significant academic influence. In addition, we found “key connectors” in the collaborative network, such as Dong Hoon Lee and Alaina M. Bever, who published relatively few papers but played an extremely important role in linking the network. They are indispensable for promoting academic exchange and cross-team collaboration.

#### Citation Analysis of Journals

The results showed that IBD nursing research is dominated by leading general medical journals, such as *The Lancet*, *New England Journal of Medicine*, *Journal of the American Medical Association*, and *British Medical Journal*, which rank among the top in total citation frequency and provide a solid evidence base for the field. Furthermore, we found that core gastroenterology journals formed a professional foundation, whereas IBD specialty journals supported in-depth development in the field. The *American Journal of Epidemiology* (intermediary centrality = 0.91) was the most important interdisciplinary knowledge integration hub in this field, connecting epidemiology, metabolism, and immunology research. The specialist journals *Gastroenterology and Inflammatory Bowel Diseases* played a dual role, combining high literature output with high intermediary centrality. In addition, the highly interdisciplinary nature of journals such as *Arthritis & Rheumatology* and *Diabetologia* revealed a deep overlap between IBD and research on autoimmune and metabolic diseases.

### Analysis of Hot Topics and Trends in Global IBD Nursing Research

#### Evolving Focus from Disease Management to Psychosocial Support and Comorbidity Care

The keyword co-occurrence results showed that nursing practice was centered on care (centrality: 0.25) and management, which formed the core of the network. In addition, quality of life ranked among the top five high-frequency keywords, indicating that it is a research hotspot and focus in the field of IBD nursing. In combination with keyword cluster analysis #12 and depression, recent research hotspots extended to the psychosocial field. Research showed that, among patients with IBD, approximately 20.9% experience impaired physical health, 13.7% experience impaired mental health, 20.0% experience pain, 18.2% experience anxiety, 15.5% experience reduced satisfaction with their social roles, 10.9% experience physical functional impairments, 10.0% experience fatigue, 7.3% experience depression, and 5.5% experience sleep disorders, all of which significantly impact the patients’ quality of life [20]. Furthermore, patients with IBD generally exhibit low levels of disease knowledge, poor adherence to treatment, and inadequate self-management behaviors [21,22]. Patients have varying levels of needs in terms of physiology, information, daily life, emotional and social support, psychological and mental health, and knowledge related to self-management and disease progression [9,23].

Based on the results of keyword co-occurrence and clustering, personalized care for pediatric patients warrants further attention. Studies have shown that approximately 25% of patients with IBD develop the condition during childhood or adolescence [24]. Adolescents with IBD have a significantly increased risk of developing anxiety or depression [23] and face greater physical, psychological, and social burdens and needs [25], warranting greater attention and support. With advancements in information technology, future research can address pediatric patients’ needs through interventions focused on the quality of life, intelligent self-management, and psychological care. Technology-based interventions can improve disease symptoms and enhance overall well-being.

The high-frequency terms “risk” (frequency: 188; centrality: 0.24), “prevalence” (165), and “health” (131) indicate a growing emphasis on preventing complications and improving patient outcomes. The association between IBD, rheumatoid arthritis, and coronary heart disease underscores the importance of comorbidity management and highlights an emerging research focus on the mechanisms underlying autoimmune-cardiovascular overlap. Research has shown that IBD frequently coexists with cardiovascular, metabolic, and neuropsychiatric diseases and malignancies during onset and progression, further complicating disease outcomes [26]. Therefore, future studies should explore IBD comorbidity care to establish IBD comorbidity risk prediction models, with the aim of preventing and managing such comorbidities and improving patient outcomes. Advancement of Research on IBD Care from Basic Mechanisms to Research Methodology, with Gradual Progression Toward Precision Medicine

The keyword “double blind” showed high intermediary centrality, reflecting the importance of evidence-based nursing. The high intermediary centrality of basic medical keywords (e.g., activation and expression) reflects the growing integration of mechanistic research with clinical nursing, and cross-disciplinary work linking molecular mechanisms, microbiology, and nursing is gradually increasing. Recent studies have introduced oral monoclonal antibody therapies that demonstrate preliminary efficacy and can be delivered via robotic pills [27]. Future research should advance self-management strategies and quality-of-life outcomes within the context of precision medicine and robotic drug delivery. Designing real-world longitudinal studies will further promote the development of nursing practice toward precision, personalization, and preventive cross-specialty integration.

### Limitations

This study primarily relied on the Web of Science database for literature retrieval. Although this is an authoritative academic literature database, it may not cover all relevant fields when collecting journals. In addition, IBD nursing research published in languages other than English were not included, which may have limited the global scope of the findings and overlooked relevant studies in specific cultural contexts. To address these limitations, future studies should focus on multiple databases such as PubMed and CINAHL. Combining these sources with methodologies such as systematic reviews could enable a more in-depth exploration of key topics related to IBD care.

## Conclusion

This study showed that research in the field of IBD care is currently stable but warrants continued attention from scholars. Patients’ quality of life, self-management, and psychological status remain popular research topics, with children and adolescents being a key population. Multidisciplinary, integrated research that leverages artificial intelligence is expected to shape future directions in this field. It is recommended that domestic scholars conduct targeted research on IBD care across diverse patient populations, especially adolescents. Efforts should focus on personalized, precise, and intelligent self-health management models or strategies based on artificial intelligence and patient-centered needs to improve service quality, alleviate patient symptoms, and enhance patients’ quality of life. Furthermore, institutions and research teams should strengthen collaboration, broaden and deepen their research, publish high-quality academic work internationally, and expand their global influence.

## Data Availability

The data that support the findings of this study are available from the authors or corresponding author upon reasonable request.

## Acknowledgments

We sincerely thank all members of the research group for their enthusiastic assistance and valuable suggestions in the topic selection, research approach, literature retrieval, data analysis, and writing of the thesis. This research was supported by the Zigong City Key Research Base for Philosophy and Social Sciences Health Humanities Research Center [project JKRWY23-22].

## Notes

### Competing Interest Statement

The authors have declared no competing interest.

### Funding Statement

Yes

### Author Declarations

This protocol has been registered prospectively in the International Prospective Register of Systematic Reviews (PROSPERO) under registration number 1156125.

